# Amniotic fluid extracellular vesicle proteome reveals fetal response to congenital cytomegalovirus infection

**DOI:** 10.64898/2026.07.21.26358423

**Authors:** Ishara Atukorala, Sally Beard, Ching-Seng Ang, Sepideh Valimehr, Luc De Catte, Natalie Hannan, Lisa Hui

## Abstract

**Introduction:** Congenital cytomegalovirus (cCMV) is the most common congenital viral infection and a leading non-genetic cause of neurodevelopmental impairment. Current diagnostic methods using fetal biofluids provide limited insight into fetal pathophysiology. Extracellular vesicles (EVs) in amniotic fluid (AF) are a promising source of stable biomolecules that reflect real-time fetal physiology. This proof-of-concept study compared amniotic fluid EV (AF-EV) characteristics in fetuses with severe CMV infection with those of uninfected fetuses and aimed to develop hypotheses about fetal response to cCMV *in utero*.

**Methods:** AF samples were collected from pregnancies with symptomatic CMV infection and gestational-age-matched uninfected controls (4 pairs, n=8 total). EVs were isolated and characterised by Western blotting, cryo-electron microscopy, and nanoparticle tracking analysis. Label-free quantitative proteomics identified CMV-associated changes in the AF-EV proteome.

**Results:** CMV-infected AF showed higher vesicle levels (Hedges’ g = 1.55), indicating inflammation and virus-induced changes in EV biogenesis. Proteomic analysis found 8.6% of proteins dysregulated. Upregulated proteins included haemoglobin subunits, immunoglobulin heavy chain mu, and myeloperoxidase (Hedges’ g = 1.51 to 1.88), indicating haemolysis and immune activation. Eleven host proteins related to neurodevelopment, mitochondrial function, lipid metabolism, and Golgi trafficking were absent in infected cases, indicating viral disruption of host pathways. Protein enrichment analysis revealed differences in neurological, haematological, and immune pathways, aligning with severe cCMV pathology.

**Conclusion:** This study acts as a proof-of-principle investigation of the AF-EV proteome in cCMV. Although the results highlight key protein signatures associated with severe fetal outcomes, they primarily serve to generate hypotheses and inform larger prospective studies.

## OBJECTIVE

Congenital cytomegalovirus (CMV) infection is the leading non-genetic cause of paediatric sensorineural hearing loss and neurodevelopmental impairment (1). Quantitative polymerase chain reaction (PCR) for CMV DNA in amniotic fluid is the diagnostic gold standard, but it provides limited insight into the severity of fetal disease, organ involvement, or pathogenesis. The species specificity of human CMV limits its modelling in animal or standard *in vitro* systems. A deeper understanding of the molecular pathophysiology of congenital CMV (cCMV) could help explain why some fetuses develop severe sequelae while others remain unaffected. Although amniotic fluid holds great potential, it remains underexplored as a source of mechanistic insight into congenital diseases in live human fetuses. Extracellular vesicles (EVs) in amniotic fluid are stable, membrane-protected carriers of proteins that reflect real-time fetal and placental physiology. We have previously characterised gestational age-related amniotic fluid EV (AF-EV) signatures in physiological pregnancies (2) and amniotic fluid cell-free RNA profiling in cCMV-affected pregnancies (3). Building on this work, we performed a proof-of-concept comparison of the AF-EV proteome in fetuses with severe symptomatic CMV infection and uninfected controls to generate hypotheses regarding the fetal molecular response to congenital CMV in utero.

## STUDY DESIGN

This exploratory case-control study used archived second-trimester amniotic fluid from 4 pregnancies with severe symptomatic CMV infection, confirmed by PCR for CMV DNA in the amniotic fluid and by placental villitis, autopsy findings, or postnatal sequelae. The CMV-negative controls were matched to cases for gestational age and fetal sex (Table 1). Samples were collected at two sites with institutional ethics approval. AF-EVs were isolated by differential centrifugation, filtration, and ultracentrifugation. They were characterised by Western blot, cryo-electron microscopy, and nanoparticle tracking analysis as per MISEV2023 guidelines (4). Label-free, data-independent acquisition mass spectrometry was performed in technical triplicate for each of the 8 samples to quantify the AF-EV proteome. Differential protein abundance between groups was assessed by Student’s t-test with permutation-based false discovery rate correction (false discovery rate 0.05, S0=0.1, 250 randomisations). Hedges’ g effect sizes were calculated to control for the bias due to small sample size. Tissue, phenotype, and Reactome pathway enrichment analyses of differentially abundant proteins were performed using the STRING database. Detailed methods are included in the Supplementary Materials.

**Table 1.** Clinical characteristics of the samples.

| Pair | Cases (amniocentesis CMV PCR positive) |  |  |  | Controls (amniocentesis CMV PCR negative) |  |  |  |  |
| --- | --- | --- | --- | --- | --- | --- | --- | --- | --- |
|  | GA at amnio (weeks) | Sex | Indication for amniocentesis | Pregnancy outcome | GA at amnio (weeks) | Sex | Indication for amniocentesis | Newborn urine CMV result | Pregnancy outcome |
| 1 | 23 | F | Abnormal ultrasound | Termination of pregnancy for severe CMV-related anomalies, placental CMV villitis | 23 | F | Maternal seroconversion (-2+2 weeks) | Negative | Term live birth |
| 2 | 22 | F | Maternal CMV seroconversion (4-8 weeks) | Termination of pregnancy for severe CMV-related anomalies, placental CMV villitis, abnormal microarray* | 21 | F | Maternal seroconversion (10-14 weeks) | Negative | Term live birth |
| 3 | 22 | F | Maternal CMV seroconversion (9-13 weeks) | Perinatal death, CMV-related sequelae confirmed on autopsy | 21 | F | Maternal seroconversion (0-4 weeks) | Positive | Asymptomatic congenital CMV in term live birth |
| 4 | 26 | M | Maternal seroconversion (4-26 weeks) | Termination of pregnancy for severe CMV-related anomalies, severe fetal growth restriction | 24 | M | Short long bones on the 5th centile | Negative | Term live birth |

GA, Gestational Age; PCR, Polymerase Chain Reaction; TOP, termination of pregnancy; F, Female; M, Male
\* Pregnancy was also associated with a genomic copy number variant

## RESULTS

AF-EVs from CMV-infected and uninfected fetuses expressed canonical EV-enriched markers (Alix, CD9, CD63) and displayed characteristic vesicle morphology by cryo-electron microscopy, with a mode diameter of approximately 140 nm (Figures A-C). CMV-infected amniotic fluid samples contained significantly higher EV concentrations than controls (Hedges’ g = 1.55), whereas the mean protein content per vesicle was unchanged (Figures D-E). Of the 2172 proteins identified with high confidence, 186 (8.6%) were significantly differentially abundant between groups (Figures F-H). Haemoglobin subunits (HBD, HBB, HBA1), immunoglobulin heavy chain mu, and myeloperoxidase were the most upregulated proteins in infected samples (Hedges’ g=1.51-1.88) (Figure I), suggesting fetal haemolysis and innate and humoral immune activation. Myeloperoxidase, a known marker of multiple sclerosis, may warrant evaluation as a candidate biomarker of disease severity for cCMV. Ten proteins were detected exclusively in uninfected controls, including regulators of Golgi trafficking (COG6, ARFGEF1), mitochondrial electron transport (UQCRC1), lipid elongation (HSD17B12), and translation initiation (EIF2B3). Loss-of-function mutations of these proteins are associated with neurodevelopmental disease, suggesting a possible mechanistic pathway for CMV pathogenesis. Enrichment analyses of the differentially expressed proteins mapped to nervous system, endocrine, urogenital, and cardiovascular tissue signatures (Figure J). These patterns recapitulate the known multi-organ tropism of severe congenital CMV. Innate immune activation, neutrophil degranulation, and alterations in vesicle-trafficking pathways were significantly upregulated in infected fetuses, indicative of an acute immune response to viral infection (Figure K). The proteins further indicated abnormalities in the nervous, head, face, musculoskeletal, and blood homeostasis (Figure L). CMV proteins were also detected on a subset of AF-EVs, consistent with prior reports of viral surface protein incorporation into host EVs (5).

**Figure.**
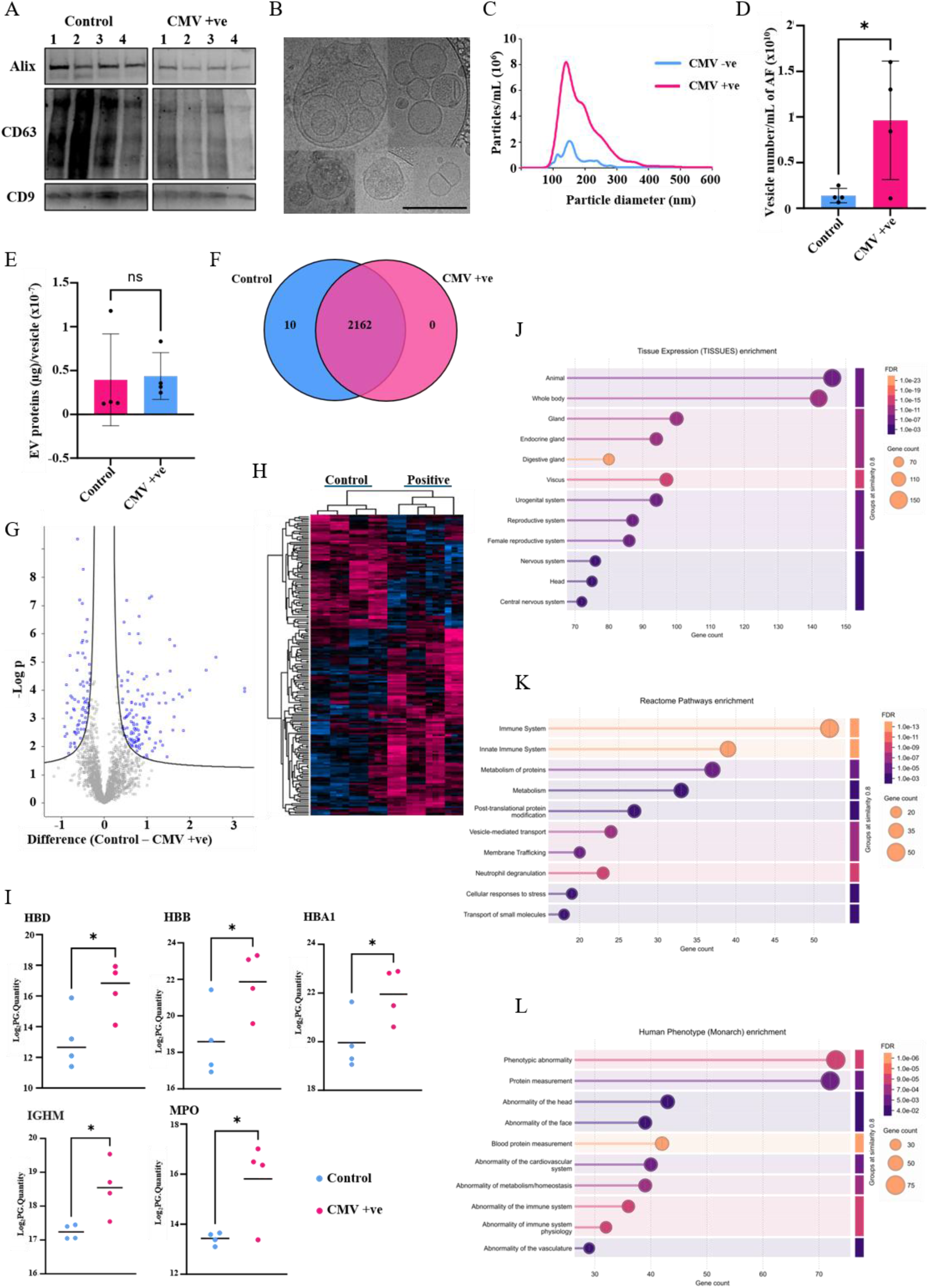
Amniotic fluid EV proteome is altered by congenital CMV infection. (A) AF-EVs were characterised by Western blot for EV markers Alix, CD63, and CD9. (B) Cryo-electron microscopy revealed typical EV morphologies (scale bar: 200 nm). (C) Nanoparticle tracking analysis showed comparable size distribution and mode diameter between groups. (D) CMV-infected samples had higher EV concentration than controls, with unchanged mean protein concentration per vesicle (E). (F) Venn diagram of the 2172 proteins identified, with 10 unique to controls. (G) Volcano plot of differentially abundant proteins (difference, control vs CMV-positive; -log p). (H) Heatmap of the 186 significantly differentially abundant proteins by unsupervised hierarchical clustering. (I) The 5 most upregulated proteins in CMV-infected AF-EVs (HBD, HBB, HBA1, IGHM, MPO). STRING DB enrichment analysis of differentially abundant proteins by tissue expression (J), Human Phenotype/Monarch terms (K), and Reactome pathways (L). Statistical comparisons used Hedge’s g (D,E,I), unpaired, 2-tailed Student t tests with permutation-based FDR correction (F-H); n=4 per group. AF-EV, amniotic fluid extracellular vesicle; CMV, cytomegalovirus; FDR, false discovery rate.

## CONCLUSION

The AF-EV proteome captures a molecular fingerprint of severe fetal CMV infection, revealing markers of haemolysis and immune activation, along with the selective loss of host neurodevelopmental, mitochondrial, and Golgi-trafficking proteins. These findings are derived from a small but well-characterised paired cohort, with technical replication and bias-corrected effect-size estimation and support the use of AF-EVs as a minimally invasive window into fetal pathophysiology that is otherwise inaccessible in vivo. Limitations include the small sample size reflecting the rarity of second-trimester amniotic fluid samples from symptomatic cCMV, available for research. Validation in larger, prospective cohorts is planned, to investigate these candidate protein signatures for biomarker development in cCMV.

## Data Availability

All data produced in the present study are available upon reasonable request to the authors. Proteomics data is available via ProteomeXchange as a partial submission with identifier PXD071698.

https://www.proteomexchange.org/

## Acknowledgements

We thank the clinical staff of University Hospitals Leuven, Leuven, Belgium, and the Department of Perinatal Medicine at the Mercy Hospital for Women, Heidelberg, Victoria, Australia, for their assistance with sample collection.

