## Supplemental Methods for "Amniotic fluid extracellular vesicle proteome reveals fetal response to congenital cytomegalovirus infection"

### Supplementary methods

#### Amniotic fluid sample collection

Amniotic fluid samples were obtained from the Mercy Hospital for Women in Melbourne, Australia and the University Hospitals Leuven in Leuven, Belgium. All samples were collected after informed consent during clinically indicated amniocentesis from November 2017 to February 2023 (Table in main text). Amniotic fluid samples were centrifuged at 300 × g for 10 minutes to remove cells and debris, then stored at -80 °C until EV isolation. This study was approved by the Human Research Ethics Committee of Mercy Health (R16-24).

#### Sample sourcing and standardised workflow

Of the sample cohort, one pair of samples was obtained from Australia and three pairs from Belgium. All samples were collected within the same gestational window and for the same clinical indications. Identical EV isolation and storage protocols were strictly followed across both sites to ensure technical consistency. All proteomics samples were processed together in a single batch; no batch effects were observed during data analysis.

#### EV isolation

AF-EVs were isolated using differential centrifugation, filtration, and ultracentrifugation. EVs were characterised according to MISEV23 guidelines (1). Amniotic fluid samples were thawed overnight at 4°C and centrifuged at 2000 g for 20 minutes at 4°C to remove cellular debris. Supernatant was centrifuged at 15,000 g for 30 minutes at 4°C, then filtered through a 0.22 µm filter. The resulting supernatant was subjected to ultracentrifugation at 120,000 g for 2 hours at 4°C to pellet small EVs. EV pellet was then washed with 0.22 µm-filtered Dulbecco's phosphate-buffered saline (DPBS; Gibco™) and centrifuged at 120,000 g for 2 hours at 4ºC. The resulting EV pellet was resuspended in filtered DPBS and stored at -80ºC until further analysis.

#### EV protein quantification

EV proteins (2 µg) of each sample immobilised on a polyacrylamide gel was fixed with fixer buffer (50% Methanol, 7% Acetic acid) and stained with Sypro Ruby gel stain (Thermo Fisher Scientific) for 24 hours. After washing the gel with wash buffer (10% methanol, 7% acetic acid), the gel was scanned using Chemidoc (BioRad) and the lanes were quantified against Benchmark$^{TM}$ protein ladder (Thermo Fisher Scientific).

**Nanoparticle Tracking Analysis**

EV size range and concentration were determined using NanoSight NS300 (Malvern Panalytical; NanoSight NTA 3.2 software). Samples were diluted 400-fold with 0.22 µm-filtered PBS and injected at an infusion rate of 50. Each sample was captured in 3 rounds of 30 seconds, with camera level and detection threshold set at 11 and 5, respectively, at 25°C.

**Western blotting**

EVs corresponding to 20 µg of protein were lysed in 4x Laemmli buffer (8% (w/v) SDS, 10% (v/v) glycerol, 200mM Tris-HCl pH 6.8, and a trace of bromophenol blue), along with 2M DTT, heated at 95°C for 2 minutes, and separated on 4-15% polyacrylamide gels (Bio-Rad). Gels were electrophoresed at 120 V for approximately 90 minutes. Proteins were transferred onto PVDF membranes (Thermo Fisher Scientific) using a wet electroblotting system (Bio-Rad), at 100 V for 1 hour, and blocked with 5% (w/v) skim milk in Tris-buffered saline with Tween 20 (TBST) for 1 hour. After washing the membranes in TBST, they were incubated with primary antibodies for Alix (Catalog # E6P9B, 1:1000, Cell Signalling), CD9 (Catalog # 10626D, 1:500, Thermo Fisher Scientific), and CD63 (Catalog # 10628D, 1:500, Thermo Fisher Scientific), at 4°C overnight. Following washing to remove excess antibody in TBST, they were incubated with the appropriate fluorescent-conjugated secondary antibodies (IRDye 680RD Goat anti-Rabbit IgG or IRDye 800CW Goat anti-Mouse IgG) (LI-COR Biosciences) at a 1:10,000 dilution for 1 hour at room temperature. After washing the membranes in TBST, they were imaged using a ChemiDoc system (Bio-Rad).

**Cryo-electron microscopy and image analysis**

Cryo-electron microscopy was used to visualise vesicles. Gold 300 mesh, lacey carbon film-coated EM grids (ProSciTech, Australia) were glow-discharged (15 mA, 30 sec) using the GloQube® Plus Glow Discharge System. A sample (4 µL of initial EV preparation) was applied onto the carbon side of the grid, which was then back-blotted for 7.0 seconds and plunge-frozen into liquid ethane using a Leica EM GP2 Automatic Plunge Freezer. The climate chamber was maintained at 90% humidity and 4°c. EM grids containing frozen samples were stored in liquid nitrogen until imaging. EVs were visualised using a Thermo Fisher Scientific Tecnai F30 electron microscope equipped with a CETA camera at the Ian Holmes Imaging Centre (The University of Melbourne). Images were recorded under low-dose conditions at 39,000x magnification with a defocus of -4 µm and a pixel size of 2.79 Å/pixel.

**Sample preparation for Mass spectrometry**

EV proteins (20 µg) were lysed in 2X lysis buffer (10% SDS, 100 mM TEAB pH 8.5). Disulphide bonds in the proteins were reduced using Tris-(2-carboxyethyl)phosphine (final concentration 5 mM) (Thermo Fisher Scientific) and alkylated with Methyl methanethiosulfonate (final concentration 20 mM). The samples were acidified to approximately 2.5% phosphoric acid. They were loaded onto S-Trap™ micro spin columns (ProtiFi, USA) with binding/wash buffer (100 mM triethylammonium bicarbonate in 90% methanol) and centrifuged at 4,000 g for 30 seconds to trap the proteins. After three washes with 150 µL binding/wash buffer, proteins were digested with Sequencing Grade Modified Trypsin (Promega) at a ratio of 1:10 (w/w) trypsin to protein for 2 hours at 47°C. Peptides were then eluted in three steps with 50 mM TEAB in water, 0.2% formic acid in water, and 50% acetonitrile in water. The eluted peptides were lyophilised using a vacuum Concentrator (Savant SpeedVac™) and reconstituted in mass spectrometry sample buffer (2% acetonitrile, 0.05% trifluoroacetic acid) to a final peptide concentration of 0.5 µg/µL.

**Data-independent acquisition (DIA) Mass spectrometry (MS)**

Liquid chromatography (LC) coupled with tandem mass spectrometry (MS/MS) was performed using an Orbitrap Ascend mass spectrometer (Thermo Fisher Scientific) equipped with a nanoflow reversed-phase high-performance liquid chromatography (HPLC) system (Ultimate 3000 RSLC, Dionex). It was fitted with an Acclaim Pepmap nano-trap column (Dionex—C18, 100 Å, 75 µm× 2 cm) and an Acclaim Pepmap RSLC analytical column (Dionex-C18, 100 Å, 75 µm× 50 cm) at the Melbourne Mass Spectrometry and Proteomics Facility (The University of Melbourne). The tryptic peptides (0.5 µg) were injected into the enrichment column at an isocratic flow rate of 5 µL/min using 2% v/v CH3CN containing 0.1% v/v formic acid for 5 minutes, after which the enrichment column was switched in line with the analytical column. The LC eluents were 5% DMSO in 0.1% v/v formic acid (solvent A) and 5% DMSO in 100% v/v CH3CN with 0.1% v/v formic acid (solvent B). The flow gradient was as follows: i) 3% B for 0-6 minutes, ii) 3-4% B for 6- 7 minutes, iii) 4-25% B for 7-82 minutes, iv) 25-40% B for 82- 86 minutes, v) 40-80% B for 86-87 minutes, vi) 80-80% B for 87- 90 minutes, and vii) 80- 3% for 90- 91 minutes. The column was re-equilibrated at 3% B for 10 minutes before the next sample injection.

For DIA experiments, full MS resolutions were set to 120,000 at m/z 200 and scanning ranged from 350 to 1400 m/z in profile mode. The Full MS Automatic Gain Control target was 250%

with an injection time (IT) of 50 ms. The AGC target value for fragment spectra was set at 2000%. Fifty windows of 13.7 Da each were used with a 1 Da overlap. Resolution was set to 30,000 and maximum IT to 55 ms. The normalised collision energy was set at 30%. All data were acquired in centroid mode using positive polarity.

**Proteomics data analysis**

DIA data were analysed using the direct DIA analysis workflow with default settings on Spectronaut® software (v. 17.5.230413.55965) against the UniProt *Homo Sapiens* database (updated Sep 2023). Trypsin specificity was set to allow two missed cleavages. Carbamidomethyl (Cys) was defined as the fixed modification, while acetylation (protein N-term) and oxidation (Met) were set as variable modifications. Pulsar search and DIA analysis results were filtered at both protein and peptide levels, with spectra matched at a false discovery rate of 1%. Precursor filtering utilised the Q value, and quantification was performed at the MS2 level. The cross-run normalisation strategy was set to automatic. Quantitative data were exported for statistical analysis into Perseus (v2.0.7.0) (26). The data were log2-transformed and grouped into two categories: CMV-infected and uninfected controls. Rows were filtered to include proteins identified in all 24 samples (8 samples x 3 technical replicates each). Ubiquitously expressed peptides present in all samples were subjected to the Student's T-test with a false discovery rate of 0.05 (multiple comparison correction) and an S0 filter set to 0.1 to identify significantly differentially abundant proteins between groups.

To identify uniquely enriched proteins in each group, we considered the presence of at least 2 technical replicates as positive and the absence in at least 2 technical replicates as negative, in each sample.

STRING DB Version 12.0, an open-source proteomic data modelling platform (2) was used to identify patterns of protein enrichment. Identified tissues/pathways/phenotypes were sorted by the number of genes in the network. Dot plots for enrichment patterns were generated using STRING DB.

**Statistical analysis**

The statistical analysis for all experiments except proteomics was conducted using GraphPad Prism 10.1.0 (GraphPad Software). We assessed data normality using the Shapiro-Wilk test. The unpaired two-tailed Student's T-test was used for parametric datasets, while the Mann-Whitney U test was applied to nonparametric datasets. Where p-values indicated statistical significance ($p<0.05$), Hedges' *g* was calculated to determine the standardised effect size. This calculation supplements p-values and accounts for the small sample size (n=4). This bias-corrected measure provides a more accurate estimate of the magnitude of the experimental effect (3). Effect size magnitudes were interpreted as small (~0.2), medium (~0.5), or large (≥0.8).
